# The VExUS System Has a Limited Clinical Impact in the Evaluation and Management of Hypoosmolar Hyponatremia: A Single-Center Observational Study

**DOI:** 10.1101/2025.02.08.25321918

**Authors:** FJ. Marques, I. Viejo-Boyano, P. González-Calero, A. Ferri, M. Gavilà, M. Mompó, P. Barrios, E. Cholbi-Vives, M. Peris-Fernandez

## Abstract

**Background:** Hyponatremia is the most frequent electrolyte disturbance in hospitalized patients, yet assessing volume status remains a diagnostic challenge. The Venous Excess Ultrasound (VExUS) system has been proposed as a noninvasive method to evaluate volume overload and guide management. However, data on its clinical impact in hypoosmolar hyponatremia are limited.

**Objective:** To investigate whether the VExUS system can support the management of acute hypoosmolar hyponatremia, detect discrepancies with physical examination in assessing volume status, and predict clinical outcomes.

**Methods:** This single-center observational study included adult inpatients with moderate or severe hypoosmolar hyponatremia (plasma sodium <=130 mEq/L). Patients underwent a standard clinical evaluation (history and physical examination) and a VExUS-based ultrasound assessment of at least two venous regions (inferior vena cava, hepatic veins, portal vein, or renal veins). Internal jugular vein measurements were used when feasible. The treating physicians of the patients were blinded to the ultrasound findings. Serum sodium levels were measured at 24, 48, and 96 hours. Treatments were based on existing clinical guidelines, and any changes in therapy were documented.

**Results:** Twenty-eight patients were enrolled, and two were excluded due to inadequate ultrasound data or new-onset cirrhosis. VExUS classification differed from clinical examination in 65.4 percent of patients (17 of 26), yet these discrepancies did not lead to significant differences in serum sodium trends or management changes at 24, 48, or 96 hours (p>0.05). Only four patients required therapeutic adjustments within 24 hours, and one displayed discrepant volume classifications. The VExUS system showed limited ability to distinguish euvolemia in patients with SIADH and demonstrated minimal correlation with laboratory parameters such as NT-proBNP and CA-125.

**Conclusions:** While ultrasound assessment is noninvasive and potentially useful, this study found that discrepancies in VExUS-based volume status classification did not significantly alter clinical outcomes or treatment strategies in hospitalized patients with hypoosmolar hyponatremia. The VExUS method should not replace standard complementary evaluations recommended by hyponatremia guidelines, although it may have a niche role when ruling in or out hypervolemia in selected cases.

## Introduction

Hyponatremia is the most frequent electrolyte disorder and affects up to 35% of hospitalized patients (1). Assessing the patient’s volume status is essential for the diagnostic and therapeutic management of hyponatremia (2). Since the development of the VExUS system (“Venous Excess Ultrasound Grading System”) aimed at predicting diuretic response in the treatment of acute kidney injury in the postoperative period of cardiac surgery (3), its use has been tested in other settings such as acute renal failure (4) and myocardial infarction (5). More recently, the use of ultrasound methods has been proposed for volume assessment in patients with hyponatremia (6), with reported usage in critically ill patients with hyponatremia (7), and during the conduct of this study for the evaluation of severe hyponatremia in hospitalized patients (8).

### Objective

The objective of this study is to assess the efficacy of VExUS in supporting the management of acute hypoosmolar hyponatremia in hospitalized patients, to verify its effectiveness in detecting discrepancies between physical examination findings and ultrasound, and to determine whether it is effective in predicting clinical outcomes in the context of hypoosmolar hyponatremia.

### Methodology

We conducted this single-center observational study in which enrolled patients were treated according to clinical guidelines for hyponatremia management (9). Within the first 24 hours after study inclusion, an ultrasound examination was performed, evaluating at least two of the following regions: inferior vena cava diameter, Doppler flow of hepatic veins, Doppler flow of the portal vein, and Doppler flow of the renal veins. As an alternative measurement, the variability of the jugular vein diameter during inspiration and expiration at a 35° incline was assessed. The physician responsible for the patient’s treatment did not have access to the ultrasound results and was therefore blinded to the sonographer’s assessment.

Different experienced clinicians performed the ultrasounds.

### Population and Sample

A sample size of 35 patients was initially planned. Adult patients diagnosed with moderate to severe asymptomatic hypoosmolar hyponatremia (plasma sodium ≤130 mEqL^-1) were included. Symptomatic patients were also included once stabilized. Exclusion criteria included any patients with pseudohyponatremia, known diabetic decompensation, uncontrolled active multiple myeloma, concurrent treatment with thiazides, hyponatremia developing in the context of urological procedures or conditions, known chronic hyponatremia, stage V chronic kidney disease or dialysis, and patients in palliative care or near end of life. Additionally, patients with known and decompensated liver cirrhosis, severe tricuspid insufficiency, and pregnant women were excluded when such conditions could affect ultrasound assessment.

Any patient for whom at least two valid ultrasound measurements could not be obtained was also excluded.

### Main Variables

The main study variable was serum sodium at 24, 48, and 96 hours after inclusion. Patients whose volume status evaluation by physical examination and VExUS differed were compared.

### Secondary Variables

Clinical data were collected on medication use and symptoms at the time of study inclusion. Physical examination data included the presence of edema, inspiratory crackles, ascites, or hepatojugular reflux. Finally, an overall clinical opinion regarding the patient’s volume status was recorded.

At the start of the study, all patients underwent testing of urinary osmolality, urinary electrolytes, and venous blood gas analysis. Patients whose blood gas analysis indicated pseudohyponatremia were excluded. It was noted whether hyponatremia was considered ADH-mediated or non-ADH-mediated based on urinary osmolality, whether results were affected by diuretic use, or whether urinary sodium was compatible with hypovolemia or hypervolemia.

During the first 48 hours, laboratory tests were performed including creatinine, estimated glomerular filtration rate, serum osmolality, LDL, HDL, triglycerides, total cholesterol, total proteins, albumin, NT-proBNP, uric acid, CA-125 (10), albumin/creatinine ratio, TSH, ACTH, and baseline cortisol.

Data on the treatment received and any changes in therapy after 24 hours of treatment or after obtaining supplementary results were recorded.

### Volume Status Assessment Using VExUS

Volume status based on VExUS was assessed according to the following principles: Hypovolemia was classified if the inferior vena cava (IVC) was <2 cm in diameter with >50% collapsibility during ventilation, along with normal venous Doppler flows.

Hypervolemia was classified if the IVC was <2 cm in diameter with <30% collapsibility, associated with any altered venous flow; if the IVC was >2 cm in diameter with <30% collapsibility and any altered venous Doppler flow; or if the IVC collapsibility was between 30% and 50% or could not be measured, associated with any degree of venous Doppler abnormality.

If normal venous Doppler flows were found and the IVC collapsibility was between 30% and 50%, or could not be determined, the situation was considered indistinguishable between euvolemia and hypovolemia.

When measurement was taken in the upper internal jugular vein, the examination was classified as suggestive of hypervolemia if the vein size increased on inspiration or did not collapse on expiration, and hypovolemia if it was collapsed on both inspiration and expiration, with a diameter reduction (anteroposterior or lateral) of >50%.

All images were saved for subsequent review.

### Statistical Analysis

Descriptive and inferential statistical analyses were performed. The different methods of volume assessment were compared with the other secondary variables. A normality test (Shapiro-Wilk) was conducted beforehand, and a non-parametric test was used when appropriate (ANOVA, Mann-Whitney, Kruskal-Wallis). A p-value <0.05 was considered statistically significant. Data analysis was carried out using R Studio, with the aid of artificial intelligence (Chat GPT 3o ®) for graphical display and inferential analysis.

### Ethical Considerations

This clinical study has been designed and conducted in strict accordance with the ethical principles set out in the Declaration of Helsinki (11). It has also been reviewed and approved by the Research Ethics Committee of Hospital La Fe in Valencia, ensuring compliance with national and international regulations on research involving human subjects.

All participants were duly informed about the study’s objectives, procedures, benefits, and possible risks, and voluntarily provided their informed consent before inclusion in the research.

## Results

A total of 28 patients were identified for inclusion during the study period. Of these, two were excluded: one for insufficient ultrasound data acquisition and another due to newly diagnosed cirrhosis decompensation. The characteristics of the included patients and the laboratory results obtained are summarized in Table 1-4. A total of 27% (n=7) of the patients had a solid organ transplant, and one patient had undergone hematopoietic stem cell transplantation.

**Table 1.** Patient Characteristics, Medications, Symptoms, and Laboratory Values.

| Variable | Total | Cases | Percentage (%) | Mean | Standard Deviation |
| --- | --- | --- | --- | --- | --- |
| Male | – | 14 | 53.84 | – | – |
| **Female** | – | 12 | 46.15 | – | – |
| **Age (years)** | 26 | – | – | 64.3 | 11.7 |

**Table 2.** Medication at Inclusion.

| **Variable** | **Total** | **Cases** | **Percentage (%)** |
| --- | --- | --- | --- |
| Fluid therapy | 26.0 | 11.0 | 42.3 |
| Thiazides | 26.0 | 0.0 | 0.0 |
| Furosemide | 26.0 | 13.0 | 50.0 |
| NSAIDs | 26.0 | 1.0 | 3.8 |
| Acetaminophen (Paracetamol) | 26.0 | 16.0 | 61.5 |
| Carbamazepine | 26.0 | 1.0 | 3.8 |
| Antipsychotics | 26.0 | 0.0 | 0.0 |
| Ifosfamide | 26.0 | 1.0 | 3.8 |
| Immunoglobulins | 26.0 | 3.0 | 11.5 |
| Levothyroxine | 26.0 | 3.0 | 11.5 |
| Corticosteroids | 26.0 | 13.0 | 50.0 |
| Recent corticosteroid withdrawal | 26.0 | 3.0 | 11.5 |

**Table 3.** Symptoms at Inclusion.

| **Variable** | **Total** | **Cases** | **Percentage (%)** |
| --- | --- | --- | --- |
| Pain | 26.0 | 9.0 | 34.6 |
| Diarrhea or vomiting | 26.0 | 10.0 | 38.5 |
| Low intake | 26.0 | 19.0 | 73.1 |
| Heart failure | 26.0 | 4.0 | 15.4 |
| Weight loss | 26.0 | 13.0 | 50.0 |
| Respiratory infection | 26.0 | 9.0 | 34.6 |
| Cancer or oncohematological patient | 26.0 | 11.0 | 42.3 |
| STUI (Lower Urinary Tract Symptoms) | 26.0 | 4.0 | 15.4 |
| Neurological symptoms | 26.0 | 2.0 | 7.7 |
| Transplant recipient | 26.0 | 8.0 | 30.8 |
| Heart (transplant type) | – | 1.0 | 3.8 |
| Lung (transplant type) | – | 5.0 | 19.2 |
| Kidney (transplant type) | – | 1.0 | 3.8 |
| Hematopoietic precursors (transplant type) | – | 1.0 | 3.8 |

**Table 4.** Laboratory Values.

| **Variable** | **Total** | **Mean** | **Standard Deviation** |
| --- | --- | --- | --- |
| pH | 26 | 7.4 | 0.1 |
| Bicarbonate (mEq/L) | 26 | 22.9 | 3.6 |
| Sodium (blood gas) (mEq/L) | 24 | 125.4 | 6.6 |
| Osm (urine) (mOsm/kg) | 25 | 342.3 | 158.0 |
| Sodium (urine) (mEq/L) | 26 | 54.3 | 38.6 |
| Potassium (urine) (mEq/L) | 26 | 29.5 | 19.1 |
| Chloride (urine) (mEq/L) | 24 | 62.0 | 43.4 |
| Urea (urine) (mg/dL) | 20 | 658.7 | 466.5 |
| FeNa (%) | 26 | 1.4 | 1.2 |
| eGFR (mL/min/1.73 m2) | 26 | 81.1 | 32.4 |
| Osm (serum) (mOsm/kg) | 23 | 268.4 | 15.1 |
| LDL (mg/dL) | 25 | 83.1 | 45.3 |
| HDL (mg/dL) | 25 | 41.4 | 25.1 |
| TG (mg/dL) | 26 | 129.2 | 97.6 |
| Cholesterol (mg/dL) | 25 | 142.4 | 58.4 |
| Total proteins (g/dL) | 23 | 5.1 | 1.4 |
| Nt-proBNP (pg/mL) | 24 | 3581.3 | 5066.9 |
| Uric acid (mg/dL) | 26 | 4.8 | 3.2 |
| Albumin (g/dL) | 26 | 3.1 | 0.7 |
| TSH (mU/L) | 25 | 2.3 | 3.4 |
| Cortisol (µg/dL) | 22 | 11.0 | 7.4 |
| ACTH (pg/mL) | 20 | 19.8 | 14.5 |
| CA125 (U/mL) | 23 | 74.2 | 51.1 |
| CAC (mg/g) | 23 | 198.6 | 353.7 |

### Main Outcome

Patients showing discrepancies in volume status evaluation by history and physical examination versus VExUS exhibited a similar clinical course, as depicted in Figure 1.

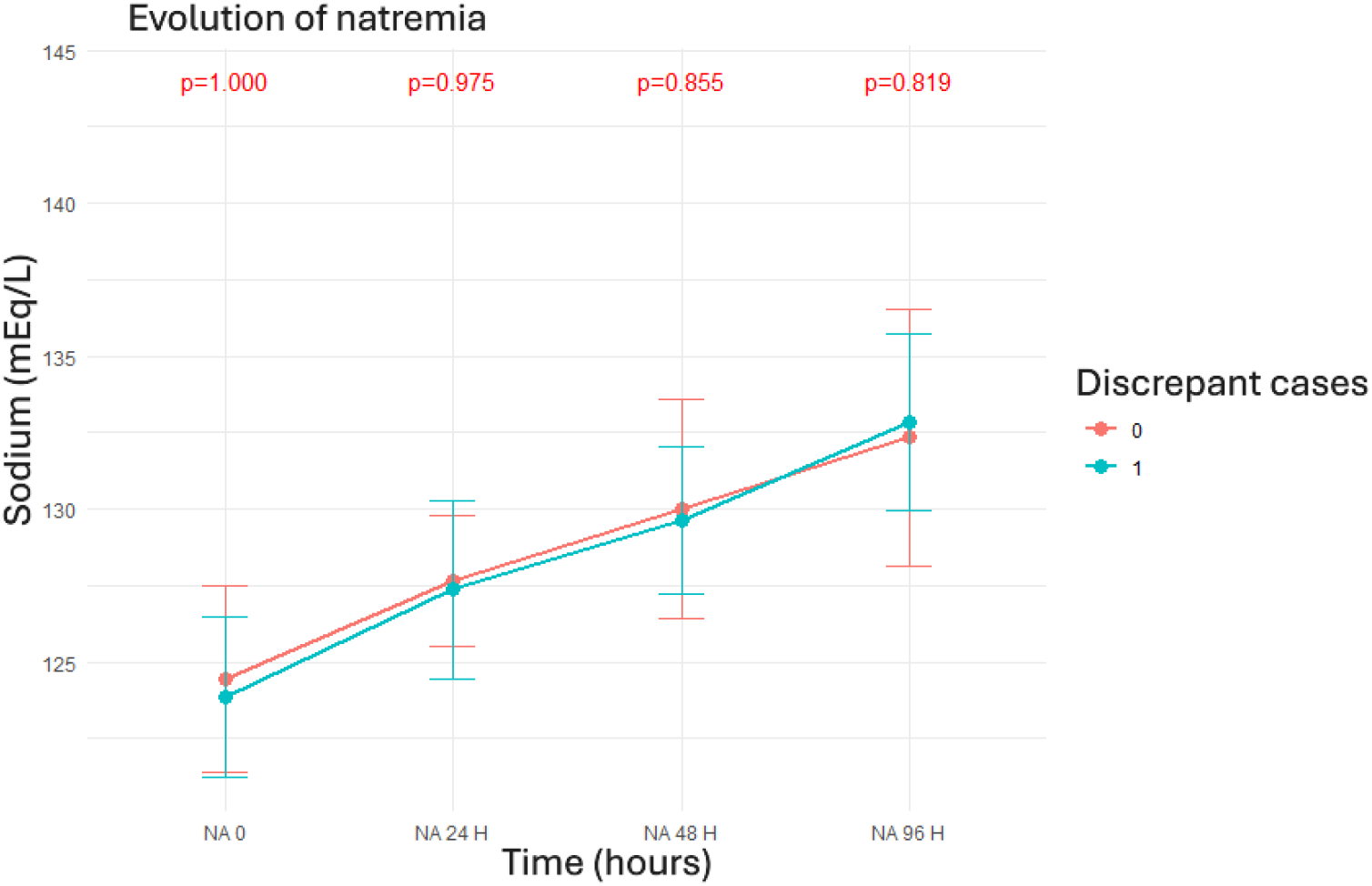

### Results of the Examination and Volume Status Assessment

According to standard methods (physical examination and history), 34% (n=9) of the patients were identified as euvolemic, 46% (n=12) as hypervolemic, and 19% (n=5) as hypovolemic based on history and physical examination. After interpreting the additional tests, 11.3% (n=3) were found to have hyponatremia not mediated by ADH. Of the ADH-mediated cases, results were compatible with SIADH in 30.8% (n=8), 38.5% (n=10) were consistent with hypovolemia or hypervolemia based on urinary sodium, and in 19.2% (n=5) of cases the results were considered uninterpretable due to diuretic use.

A total of 61.53% (n=16) of the ultrasound examinations were performed by the same examiner. The number of ultrasound measurements that could not be obtained in each region was 34% (n=9) for the IVC, 15.4% (n=4) for the hepatic veins, 3.4% (n=1) for the portal vein, and 30.8% (n=8) for the renal veins. Measurements were obtained in the internal jugular vein in 42.3% (n=11) of patients.

According to VExUS, 26.9% (n=7) of the patients were classified as euvolemic/hypovolemic, 46.2% (n=12) as hypovolemic, and 26.6% (n=7) as hypervolemic. A total of 65.4% (n=17) of patients showed discrepancies between volume status assessment by history and physical examination versus the ultrasound technique. Specifically, 9 patients were classified as hypervolemic by physical examination but as hypovolemic (n=5) or euvolemic/hypovolemic (n=4) by ultrasound. One patient was identified as hypovolemic and subsequently as hypervolemic by ultrasound, and 7 patients classified as euvolemic by physical examination were categorized as hypervolemic (n=3) or hypovolemic (n=4) by ultrasound.

Serum sodium trends based on these discrepancies were similar between groups, and the differences were not statistically significant (ANOVA; sodium at 24 hours p=0.142, 48 hours p=0.775, 96 hours p=0.667). Data are shown in Figure 2 (next).

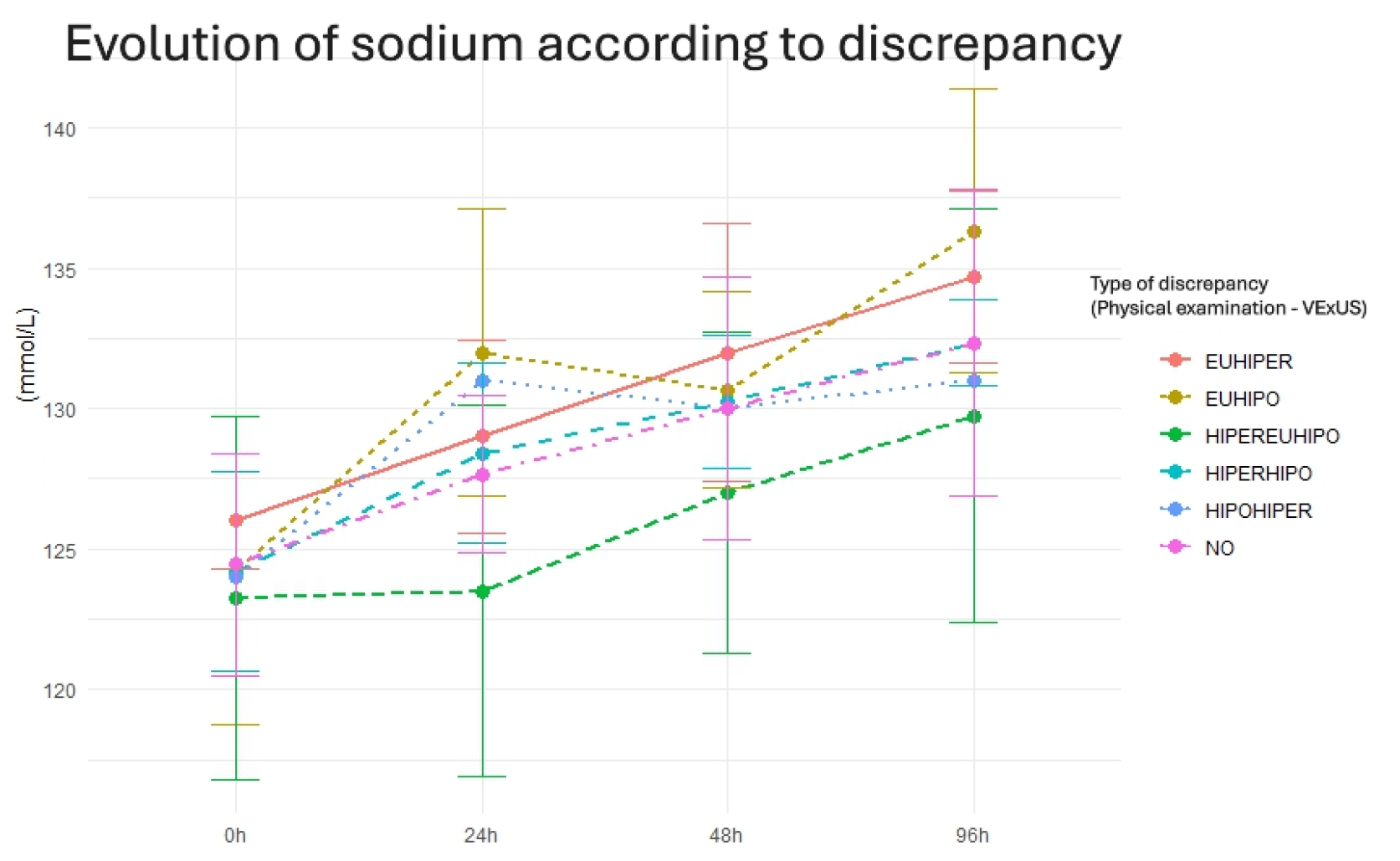

### Treatments

A total of 65.38% (n=17) were treated by recommending fluid restriction, and 30.77% (n=8) received fluid therapy. Among those who received diuretics, furosemide (26.92%) was the most commonly used, followed by tolvaptan (15.38%). A total of 7.69% (n=2) of patients received urea as initial treatment. A total of 23.08% (n=6) of patients received hypertonic saline as initial treatment, and after 24 hours 19.23% (n=5) of patients continued receiving it.

Two patients experienced early deterioration, and in both cases an infusion of 3% saline was started. As part of their treatment, cotrimoxazole was withdrawn in one patient as a possible cause of hyponatremia; in another patient, fluids were stopped, and in another, spironolactone was discontinued. One patient was found to have tacrolimus toxicity, and tacrolimus dosing had to be adjusted. Another patient had levothyroxine reintroduced after it had been discontinued by mistake at admission.

Patients who presented discrepancies between physical and ultrasound examination findings were treated similarly. Only 4 patients required a treatment adjustment 24 hours after the study began, of whom only 1 showed discrepancies in volume status evaluation.

### Comparison Between Volume Status Assessments and Surrogate Laboratory Parameters of Volume

No greater discrimination of volume status was found when comparing BNP levels according to VExUS versus physical examination, as distribution differences are similar with VExUS (Kruskal-Wallis, p=0.37) and physical examination (Kruskal-Wallis, p=0.36). However, a greater tendency toward correct identification of patients with very high BNP was observed, as shown in Figures 3 and 4. CA-125 is similarly distributed across the different volume status groups according to VExUS, but its grouping is more consistent with physical examination, albeit not statistically significant, as displayed in Figures 5 and 6.

According to VExUS, 3 out of 8 patients with SIADH were classified as euvolemic, with the rest classified as hypovolemic or hypervolemic. Of the patients who showed low urinary sodium (reflecting either hypervolemia or hypovolemia), 50% (5/10) were classified as hypervolemic, 30% (3/10) as hypovolemic, and 20% (2/10) as euvolemic/hypovolemic. All 3 patients with ADH-independent hyponatremia were classified as hypovolemic in every instance. The concordance between physical examination and interpretation of additional test results was poor (Fisher’s exact test, p=0.30).

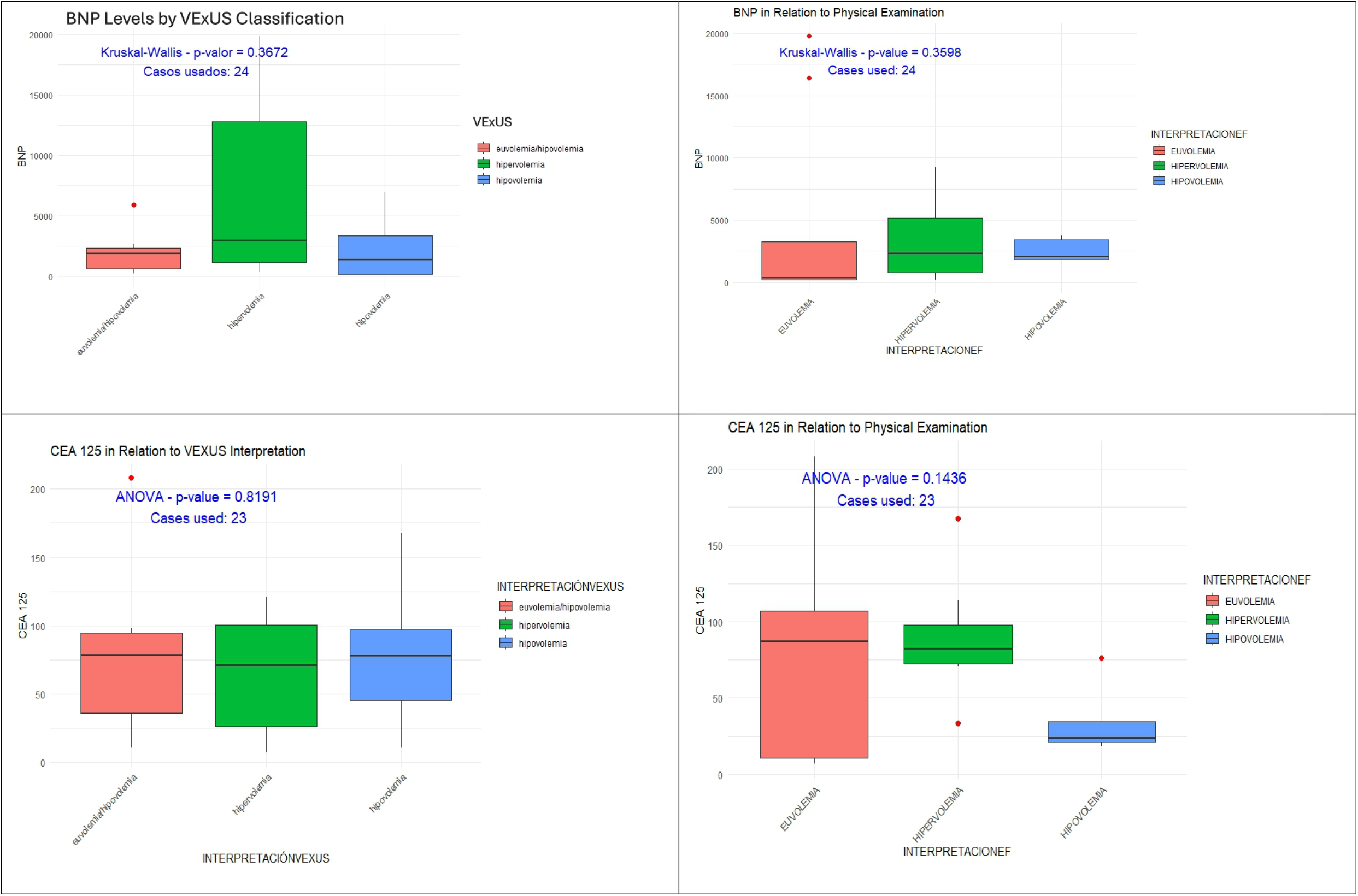

## Discussion

To our knowledge, this is the second study (8) to examine ultrasound measurements for volume status assessment in hospitalized patients with hyponatremia and the first to evaluate the clinical impact of discrepancies found.

There are significant methodological differences between this study and the one by Rahman et al. (2024). Rahman et al. used a more global examination, reviewing the presence of pleural effusion or ascites and exclusively assessing the inferior vena cava. The present study adheres to the evaluation proposed by Beaubien-Souligny et al. (2020). Furthermore, patients with moderate hyponatremia were not excluded in our study, whereas they comprise the majority of patients in this study. Rahman et al. (2024) did not exclude patients with tricuspid regurgitation or cirrhosis, a patient group that often presents with hypervolemia and hyponatremia.

The main difference lies in the primary objective. Since there is no standardized method for volume status assessment, our study focused on serum sodium trends and the need for changes in treatment to determine whether a discrepant or discordant volume classification has a significant clinical impact. Rahman et al. (2024) did not address this question.

In the present study, discrepancies in volume classification based on clinical assessment and the VExUS system did not predict worse clinical outcomes or a greater need for treatment changes.

Although there is a tendency toward poorer outcomes, particularly in patients classified as hypervolemic by clinical criteria but euvolemic/hypovolemic by VExUS, these patients met SIADH criteria and were treated with diuretics. We therefore consider that the discrepancy does not lead to different clinical management and is, hence, not clinically relevant.

A trend toward better classification of patients with high or very high NT-proBNP using VExUS was observed, though this was not the case with other surrogates like CA-125.

Patients with SIADH were classified into different volume categories by VExUS, suggesting that the technique may not accurately classify euvolemia in SIADH. This limits its applicability in a context with a high prevalence of SIADH. In our study, only 5 patients showed severe VExUS abnormalities, which may have limited its utility compared to Rahman et al. (2024).

Our study has multiple limitations. We did not reach the planned sample size during the study period, limiting the power of our inferential analyses. Most of the ultrasound examinations were performed by the same operator, which may introduce bias. Additionally, since patients were hospitalized for various reasons and managed by different specialists, the therapeutic strategy was not exclusively under the control of the study investigators, and not all changes were fully tracked. In many cases, correcting hyponatremia was not the primary priority, so some therapeutic changes were influenced by other clinical variables not thoroughly collected. However, the study reflects the general experience of a large hospital over a 10-month period, consistent with daily clinical practice.

The findings suggest a limited role for VExUS in hyponatremia for several reasons. First, different volume classifications converge on similar treatments (e.g., diuretic use in both SIADH and heart failure). In some clinical scenarios, the use of hypertonic saline is indicated, so an incorrect assessment of hyponatremia etiology may have minimal impact on the initial stages of treatment.

The ultrasound method used does not seem to effectively differentiate euvolemia from hypovolemia, as it failed to classify SIADH patients as euvolemic, and it also labeled 20% (2/10) of patients with low urinary sodium (indicating hypo- or hypervolemia) as possibly euvolemic. Correlation with other laboratory volume surrogates was poor. Therefore, its broad applicability is limited when excluding patients with heart failure or cirrhosis.

Only 5 cases of hypervolemia by VExUS were identified. The clinical context in which it might be most useful (classifying a truly hypervolemic patient as hypovolemic) occurred only once. Even so, the patient had a favorable outcome. This also aligns with the use of other ultrasound views beyond the major veins, as in Rahman et al. (2024). Neither a euvolemic nor a hypovolemic patient would present with pleural effusion or ascites, so there is no way to differentiate them on that basis.

Although ultrasound is harmless, inexpensive, and can be helpful in certain circumstances, obtaining VExUS parameters can be time-consuming (including transporting the ultrasound machine or the patient, and difficulty acquiring images in obese patients or in poor acoustic windows). Therefore, we do not consider its generalized use to be cost-effective. Notably, it was not possible to obtain an image from at least one region in 34% of cases.

Finally, given its limitations, VExUS does not eliminate the need for other complementary examinations recommended in hyponatremia clinical guidelines.

## Conclusions

In this study, discrepancies between volume assessment via the VExUS system and conventional clinical evaluation did not translate into significant or clinically relevant differences in serum sodium trends or treatment adjustments. The ability of VExUS to adequately distinguish between euvolemia and hypovolemia—particularly in SIADH patients—was limited. The time required and the frequency with which ultrasound images could not be obtained pose further restrictions to its widespread applicability. Consequently, we do not recommend that clinicians rely on VExUS findings to exclude other complementary evaluations recommended in hyponatremia management guidelines. While larger studies are needed to more precisely define its utility, the experience presented here suggests limiting the use of VExUS to clinical situations where the primary question is to confirm or rule out hypervolemia.

## Data Availability

All data produced in the present study are available upon reasonable request to the authors

## Acknowledgments

We would like to thank the Clinical Analysis Department of the Hospital Universitari i Politècnic La Fe for facilitating the conduct of this study.

